# Linking Profiles of Pathway Activation with Clinical Motor Improvements – a Retrospective Computational Study

**DOI:** 10.1101/2021.10.08.21264743

**Authors:** Konstantin Butenko, Ningfei Li, Clemens Neudorfer, Jan Roediger, Andreas Horn, Gregor R. Wenzel, Hazem Eldebakey, Andrea A. Kühn, Martin Reich, Jens Volkmann, Ursula van Rienen

## Abstract

Deep brain stimulation (DBS) is an established therapy for patients with Parkinson’s disease. *In silico* computer models for DBS allow to pre-select a set of potentially optimal stimulation parameters. If efficacious, they could further carry insight into the mechanism of action of DBS and foster the development of more efficient stimulation approaches. In recent years, the focus has shifted towards DBS-induced firing in myelinated axons, deemed particularly relevant for the external modulation of neural activity. We use the concept of pathway activation modeling, which incorporates advanced volume conductor models and anatomically authentic fiber trajectories to estimate DBS-induced action potential initiation in anatomically plausible pathways that traverse in close proximity to targeted nuclei. We apply the method on a retrospective dataset with the aim of providing a model-based prediction of clinical improvement following DBS (as measured by the motor part of the Unified Parkinson’s Disease Rating Scale). Based on differences in outcome and activation rates for two DBS protocols in a training cohort, we compute a theoretical 100% improvement profile and enhance it by analyzing the importance of profile matching for individual pathways. Finally, we validate the performance of our profile-based predictive model in a test cohort. As a result, we demonstrate the clinical utility of pathway activation modeling in the context of motor symptom alleviation in Parkinson’s patients treated with DBS.

## Introduction

Deep brain stimulation (DBS) is an effective treatment for various neurological and mental disorders, and it has become an established therapy for patients suffering from therapy-refractory Parkinson’s disease (PD). During DBS, short high-frequency pulses are delivered to subcortical brain structures via implanted electrodes. These electrodes usually have 4 or 8 contacts, each of which can be used as a current source. Modern DBS systems allow great flexibility in pulse modulation, including adjustment of width, amplitude, and frequency. Determining an optimal stimulation protocol in such a large parameter space is challenging, and to assist medical professionals in this procedure, *in silico* computational models for DBS could be of use. Besides, such models could provide insights into the action mechanism of the treatment, which in turn could drive the development of more efficient and effective stimulation paradigms.

In the basal ganglia, DBS has been hypothesized to create an “informational lesion” partly facilitated by electrical stimulation of highly excitable myelinated axons [21] that form projections across DBS targets, such as the subthalamic nucleus (STN) and the globus pallidus internus (GPi). To quantify the effect, different concepts have been proposed, among which the volume of tissue activated [8] or its approximations [13] are most commonly employed. Multiple studies have investigated correlations of symptom alleviation with voxel metrics defined based on the volume of tissue activated and its interaction with structural and functional connectivity [28, 29, 38, 43]. More recently, the concept of pathway activation modeling was proposed [23] that comprises advanced volume conductor models and anatomically authentic fiber trajectories to estimate a DBS-induced action potential initiation along pathways residing in the vicinity of the stimulation targets. To date, studies on Tourette syndrome, obsessive-compulsive disorder, treatment-resistant depression, and PD have successfully employed pathway activation modeling to predict symptom alleviation in patients [20, 24, 30, 34]. However, to our best knowledge, the method has not been applied on a cohort level to investigate correlations between DBS-induced axonal activation and alleviation of aggregated motor symptoms in PD.

In this retrospective computational study, we apply pathway activation modeling to identify the network correlates underlying the improvement of the Unified Parkinson’s Disease Rating Scale III score (motor examination, further referred to as UPRDS-III) and the Movement Disorder Society (MDS) sponsored revision of UPRDS-III in patients suffering from therapy-refractory PD. We base this prediction on pathway activation profiles defined by multiple pathways recruited by Subthalamic Nucleus Deep Brain Stimulation (STN-DBS). Correlating symptom alleviation with profiles instead of activation in individual pathways can be a more robust metric considering compensatory and adverse effects of fiber recruitment in the vicinity of the STN. Based on interprotocol scores, defined as the difference of two DBS-on UPDRS-III scores assessed in the training cohort for each patient, we construct a theoretical 100% UPDRS-III improvement profile and enhance it by analyzing the significance of activation levels in individual pathways. The performance of the resulting profile-based predictive model is then successfully tested in an independent cohort.

## Materials and Methods

### Patient Cohorts and Imaging

Two cohorts from independent DBS centers, namely Charité – Universitätsmedizin Berlin and Würzburg University Hospital, were retrospectively analyzed for derivation and validation of a pathway activation-based predictive model. The cohorts were formed based on the following criteria:

- availability of medical imaging data, such as preoperative T1-weighted magnetic resonance imaging (MRI) and postoperative computed tomography (CT), necessary for electrode localization and patient-specific modeling;
- current-controlled stimulation mode, which allows to compensate for a voltage drop on the electrode-tissue interface, reducing the computational model complexity;
- placement of the electrode inside the STN with a minimal penetration of the pallidus. The former restriction is imposed since only activation in pathways in the vicinity of the STN was investigated, and the latter criterion was applied to reduce effects of the pallidal lesioning.

As a result, 15 patients for the training (Berlin) and 19 for the test (Würzburg) cohort were admitted. Although the first cohort contained fewer patients, it provided a total of 30 datapoints: two stimulation protocols were documented for each patient as a part of another study on an algorithm-guided DBS-programming [53]. Later, this aspect was used to derive the predictive model. In both cohorts, the patients constituted a representative sampling of a clinical PD-DBS cohort (see Table 1). All received octopolar DBS electrodes bilaterally to STN (Fig. 1), either using omnidirectional or directional DBS electrodes (Boston Scientific Vercise™, Marlborough, MA, USA). Stimulation was performed using a conventional current-controlled DBS signal: a rectangular pulse of 20–60 *µs* length, 79–185 *Hz* repetition rate, each followed by an extended charge balancing period at low amplitude. Motor performance in the training and the test cohorts was evaluated with either UPDRS-III or MDS-UPDRS-III, respectively. These scores are strongly (*r >* 0.95) correlated [42], and, for brevity, both will be referred to as UPRDS-III. They were taken at baseline (off medication, DBS-off) preoperatively in the test cohort, and at least 6 months after surgery (same day as DBS-on scores) in the training cohort. UPDRS-III scores under active DBS (off medication) were acquired at least after 6 months in both cohorts.

**Table 1.** Cohort Demographics and Clinical Outcomes.

| Cohort | No. (female) | Age, years | Disease duration, years | MDS-UPDRS-III<br>Pre-op baseline | UPDRS-III<br>Post-op baseline | (MDS-)UPDRS-III<br>DBS-on Med-off |
| --- | --- | --- | --- | --- | --- | --- |
| Training | 15 (2) | $62.4 \pm 6.2$ | $12.8 \pm 4.8$ | — | $40.6 \pm 8.8$ | $23.3 \pm 8.7$ |
| $P_1$ | —//— | —//— | —//— | — | —//— | $21.5 \pm 8.8$ |
| $P_2$ | —//— | —//— | —//— | — | —//— | $25.1 \pm 8.3$ |
| Test | 19 (7) | $55.5 \pm 5.7$ | $9.8 \pm 3.4$ | $43.3 \pm 9.4$ | — | $22.3 \pm 9.6$ |

Mean values ± standard deviation are reported. In the test cohort, baseline (off medication, DBS-off) was assessed preoperatively and in the training cohort at least 6 months after surgery. The training cohort contains information on two stimulation protocols ( $P_1$ and $P_2$ ) for each patient; their score difference over the cohort is $3.6 \pm 7.3$ . (MDS-)UPDRS-III – (Movement Disorder Society sponsored revision of) Unified Parkinson Disease Rating Scale, motor examination.

**Figure 1.**
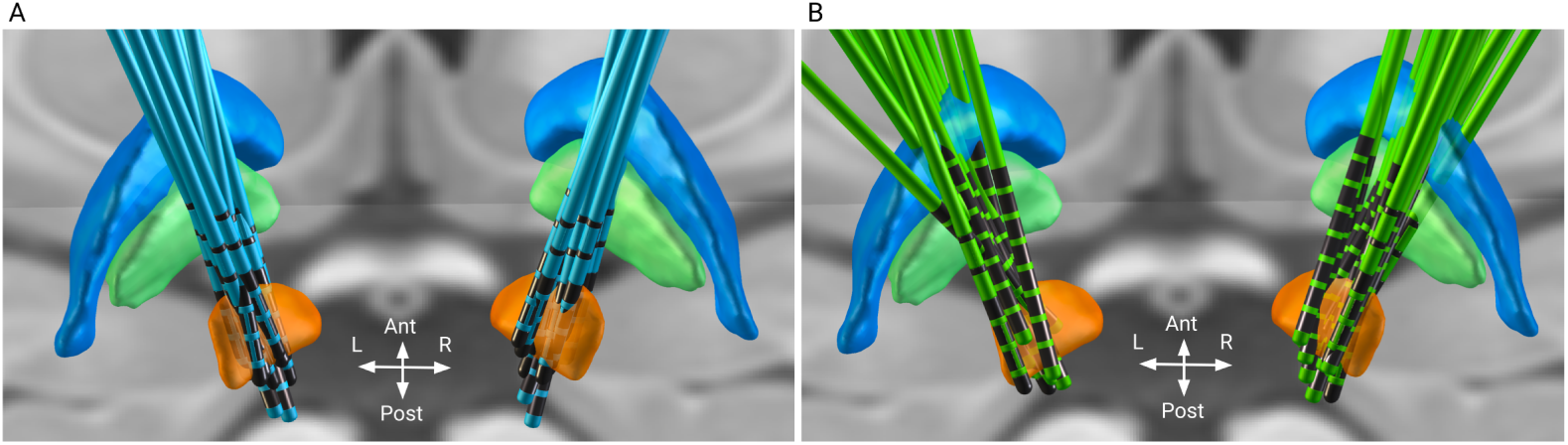
Reconstruction of DBS electrodes in the training (Berlin, A) and the test (Würzburg, B) cohorts. Electrodes are visualized in standard stereotactic Montreal Neurological Institute (MNI) space using Lead-DBS. Displayed brain structures are defined by the DISTAL atlas [15] and include subthalamic nucleus (orange), globus pallidus externus (blue) and globus pallidus internus (green).

**Figure 2.**
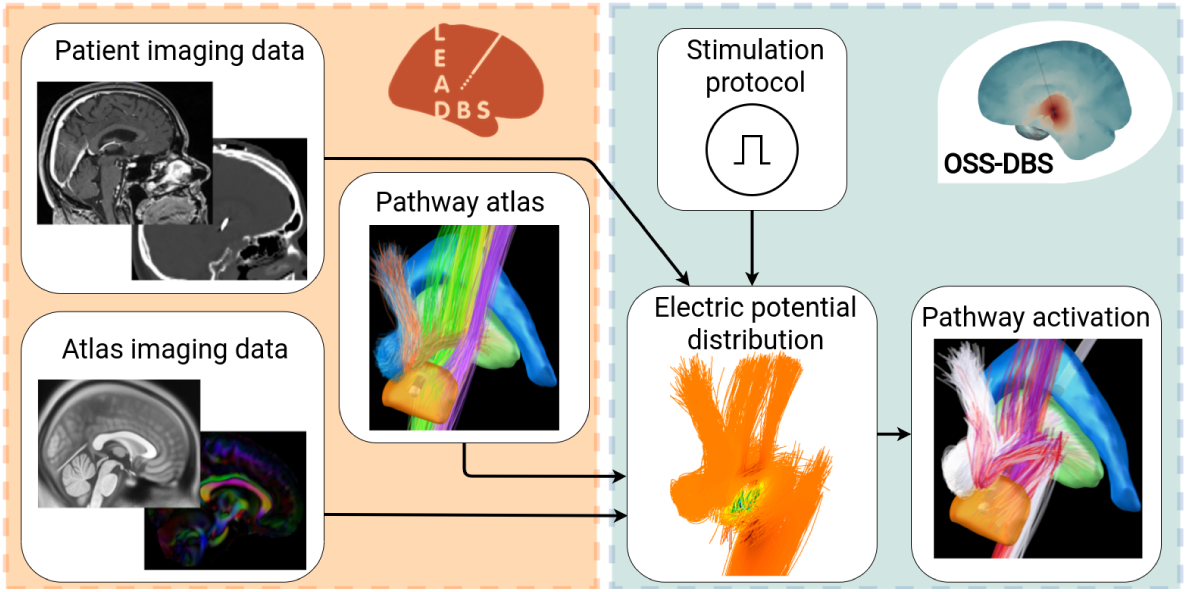
Dataflow for computing pathway activation. Based on patient imaging and brain atlases, Lead-DBS (orange box) reconstructs the electrode and provides a description of tissueand water diffusion distributions in the brain. These data are used by OSS-DBS (green box) to create an accurate patient-specific volume conductor model. The model is then employed to compute electric potential distribution in space and time along axon models allocated on trajectories described by a pathway atlas. Finally, for the given distribution, the cable equation is solved to probe axonal activation, i.e., occurrence of an action potential in response to DBS.

Imaging data was processed using Lead-DBS software [26, 27] (lead-dbs.org). Postoperative volumes and preoperative weighted multimodal MRI scans were linearly co-registered using SPM12 [16] (fil.ion.ucl.ac.uk/spm). This was followed by a non-linear normalization step of co-registered patient scans to stereotactic Montreal Neurological Institute (MNI) space using Advanced Normalization Tools (ANTs, stnava.github.io/ANTs/) ’SyS’ algorithm [5]. We also accounted for potential non-linear displacements introduced by brain shift using an additional refinement of the co-registration step that focused on the subcortex. Electrodes were localized based on CT scans using PaCER [32], and results were visually evaluated and refined, if necessary.

Bioelectrical effects of DBS were modeled in patient-specific (native) space. For that, patient-specific brain tissue segmentations (grey matter, white matter and cerebrospinal fluid) were obtained based on tissue probabilistic mapping [16] derived from the multispectral MNI template and T1-weighted MRI images. To account for tissue anisotropy, the mean diffusion tensor data of the human brain [56] were transformed into patient-specific space using the inverse deformation field derived from the diffeomorphic normalization procedure. The same procedure was applied to a basal ganglia pathway atlas [47] that describes fiber distribution and classification necessary for pathway activation modeling. Created under the guidance of expert neuroanatomists, it provides an accurate description of axonal trajectories affected by STN stimulation.

### Field and Pathway Activation Modeling

Isotropic conductivity values for grey and white matter were evaluated in the frequency domain based on [17], but omitting *α*-dispersion as proposed in [57] and additionally upscaled. The conductivity *σ* of non-dispersive cerebrospinal fluid was fixed to 2.0 *S/m*. In the power spectrum of a conventional DBS signal (up to 1 *MHz*), the conductivities were monotonically increasing (*σ*_*grey*_: 0.168–0.235 *S/m* and *σ*_*white*_: 0.120–0.153 *S/m*). To account for this dispersion, the Fourier Finite Element Method [8] was applied to solve the quasistatic formulation of Maxwell’s equations that describes the spatial distribution of the electric potential *φ*(**r**):

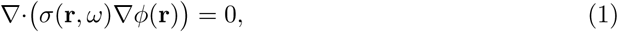

where *ω* = 2*πf* is the angular frequency of one of the harmonics that compose the DBS signal. Capacitive properties of brain tissue can be neglected in Eq. 1 due to its relatively low contribution after omitting *α*-dispersion. An octave band approximation method [6] was used to reduce the number of computations in the frequency domain. The solution in the time domain is then retrieved using an Inverse Fourier Transform. Anisotropy, especially prominent in white matter tracts [18], was modeled by expressing conductivity in terms of tensors defined according to the mean diffusion tensor data that were normalized voxel-wise following the volume conservation approach [22] and scaled by the isotropic conductivity of brain tissue. The electrode-tissue interface was neglected assuming its minor effect on current-controlled stimulation [8] with a charge density per phase below 0.03 *mC/cm*^2^. Nevertheless, the electrode’s encapsulation layer was accounted for by removing axons within a 0.1 *mm* vicinity, where neuron degeneration and glial scarring occur. The accuracy of the Finite Element Method computations was controlled based on the convergence of the electric field and the current, and elements with large deviations were refined.

The obtained distribution the of extracellular potential in space and time was used to solve a double cable equation of a myelinated axon model described in [41]. The models were allocated on the trajectories delineated in [47] (for passing fibers, the closest point on the trajectory to active contacts was treated as the midpoint seed), and the pathway activation rate was computed as a fraction of axons in the pathway that elicited an action potential in response to the DBS pulse. In the present study, we modeled activation in the corticofugal and the hyperdirect pathways (HDP) originating in the primary and premotor cortex, as well as the supplementary motor area. To reduce the computational costs, the corticofugal pathway was uniformly downsampled from 5000 to 1250 streamlines. In addition, activation in the pallidosubthalamic pathways (sensorimotor portion) and the pallidothalamic pathways (ansa lenticularis and lenticular fasciculus) was investigated. Besides, we computed the extent of direct DBS recruitment of the passing cerebellothalamic tract associated with tremor suppression [10]. In total, 4000 axon models of 16 pathways per hemisphere were deployed for pathway activation modeling, with some axons being later removed due to their intersection with the electrode, the encapsulation layer or cerebrospinal fluid.

In the present study, the fiber diameters and the number of nodes of Ranvier, which together defined the axonal length, were fixed for axons within one pathway. A fiber diameter of 3.0 *µm* was chosen for axons of the local STN-GPe and STN-GPi pathways with a length of 10 *mm* and 6 *mm*, respectively (35 nodes and 21 nodes of Ranvier). For other pathways, the fiber diameter was set to 5.7 *µm*, which is on a larger side for data reported in [40]. The same axon model was employed in [31], where it was noted that larger fiber diameters improve predictability of evoked potentials. In a preliminary analysis, we also tested a 12.0 *µm* fiber diameter as suggested by the authors, but this setup predicted a high activation in the corticofugal pathway, which is unlikely for clinically accepted protocols due to evoked motor contractions. For computational reasons, the length of the axons of passage and the HDP was limited to 20 *mm* (40 nodes of Ranvier), thus not covering the whole length of the corresponding projections. Nevertheless, this truncation is acceptable due to a minor effect of DBS on distant axonal compartments, and a preliminary analysis with longer axons yielded the same activation rates.

All steps described in this subsection were carried out using the open-source simulation software OSS-DBS [7] that was developed for highly automated DBS modeling in heterogeneous anisotropic and dispersive volume conductor models. For the present study, the software was implemented as a computational backend, freely distributed along with Lead-DBS. The coupling allowed a seamless transition between state-of-the-art processing of medical imaging and accurate modeling of DBS-induced electric field with subsequent quantification of its effect on neural tissue using cable models. The OSS-DBS computations were encapsulated in Docker containers (docker.com/) deployed on Intel Xeon(R) Gold 6136 CPU @ 3.00 GHz x 48 cores with 376.6 GB of memory.

### Analysis of Pathway Activation Profiles

As a first exploratory step, we calculated correlations between activation rates of individual pathways and the UPDRS-III improvement from baseline in the training cohort. We employed the Pearson correlation coefficient, assuming a linear interaction between the quantities. The Spearman rank correlation was comparable for all tests presented in this study.

Secondly, we analyzed activation rate profiles **AR** (vector quantities composed of activation rates over all 16 pathways) and their correlation with alleviation of motor symptoms. Since the training cohort contained two DBS-on datapoints per patient, we decided to use these measurements to derive a vector of the interprotocol UPDRS-III improvement in the pathway activation space. The difference in UPDRS-III scores between two protocols allowed us to compute a distance to a theoretical pathway activation profile of 100% improvement, i.e., an optimal activation rate profile:

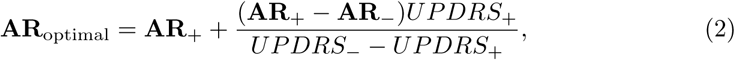

where subscripts “+” and ” − “refer to the better and the worse performing stimulation protocols, respectively. Such a definition of the optimal pathway activation profile is preferable over derivations using baseline, where only endogenous activity is present that is not quantified in the model and assumed to be overwritten by the DBS-induced activation of fiber tracts.

The difference between two profiles was quantified with the Canberra distance [36]

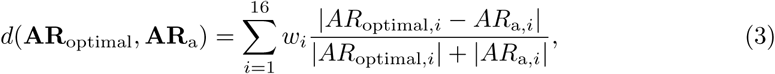

where *w*_*i*_ is the weighting factor for the *i* th pathway, whose default value is 1.0. The Canberra distance was chosen based on a preliminary analysis within the training cohort where it showed the highest predictive ability in comparison with other metrics, such as Bray-Curtis dissimilarity and Euclidean distance. It is noteworthy that this normalized metric demonstrated the best performance, thus sparing us the argument of whether activation rates (percent of fibers present in the tract) or absolute activation (amount of fibers activated in each tract) should be employed when analyzing pathway activation results.

The next problem was to determine which patients, i.e., pairs of datapoints, to use to derive **AR**_optimal_. At first glance, the patient with the best improvement between two DBS-on protocols would be a good candidate. However, the best improvement does not guarantee the shortest path to the optimal activation rate profile. Instead, we considered patients who had the highest interprotocol UPDRS-III improvement normalized by the mean of the difference vector (**AR**_+_ − **AR**_−_). To increase the robustness of the predictive model, **AR**_optimal_ can be averaged among several patients. In the present study, we picked three patients with a prominently higher normalized UPDRS-III improvement. Additionally, we tested the predictive ability of **AR**_optimal_ derived from all datapoints in the training cohort using a “leave-one-out” approach.

Previous experimental and clinical studies showed that stimulation of specific pathways in the vicinity of the STN associated with symptom alleviation and occurrence of side-effects [9, 10, 48, 51]. Therefore, we presume that the importance of matching the optimal profile is not uniform among pathways. To test this hypothesis, we optimized the weighting factor *w*_*i*_ for each pathway using the training cohort, but excluding those recruited for **AR**_optimal_. The optimization was conducted according to the following procedure. First, a patient was excluded, and the rest were randomly shuffled and split. Next, for one set, the weighting factors were optimized to maximize the inverse correlation of the Canberra distance and the UPDRS-III improvement, both from baseline and between protocols. The second set was then used to test the predictive ability of the weighted metric. The procedure was repeated for all patients. Removal of a patient and random shuffling was employed to estimate convergence of the optimal weighting factors across the cohort, and multiple trials yielded no significant disparities introduced by the shuffling. Note that the limited number of datapoints overall necessitated use of the same patients (but in different combinations) in both sets.

## Results

### Pathway Activation and UPDRS-III Improvement

Pathway activation rates computed for both cohorts are presented in Fig. 3. Analysis of activation in individual pathways and UPDRS-III improvement from baseline revealed only one pathway with a statistically significant positive correlation, namely, the HDP branch to the face-neck region of the primary motor cortex. However, it was not predictive when later applied to the test cohort. Additionally, we observed that increase in activation in the cerebellothalamic pathway correlated with interprotocol UPDRS improvement of tremor (see Suppl. 1). Interestingly, there was no correlation with the UPDRS-III improvement from baseline, possibly due to irrelevance of the metric activation for akinetic-rigid patients and contribution of the subthalamic projections to the tremor alleviation. The weak correlations for individual pathways might indicate an interplay of activation in different pathways.

**Figure 3.**
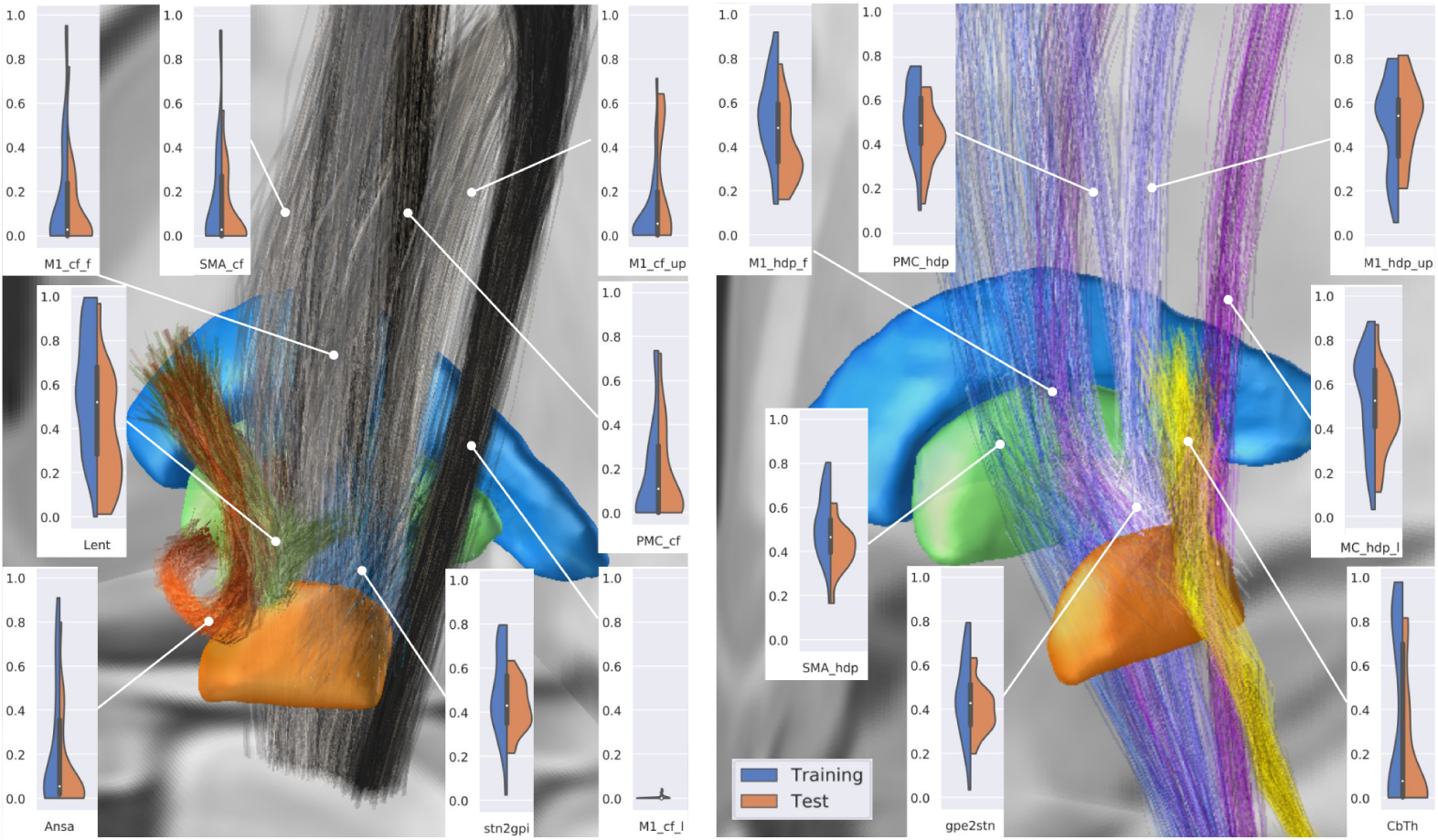
Violin plots of pathway activation rates across both cohorts (two stimulation protocols for each patient in the training cohort) visualized with the correspnding pathways in Lead-DBS in MNI space. MC and PMC refer to the primary and premotor cortical regions, SMA – supplementary motor area; cf and hdp are the corticofugal and the hyperdirect pathways, respectively; Ansa – ansa lenticularis, Lent – lenticular fasciculus, CbTh – cerebellothalamic pathway; l, f, up - lower extremity, face-neck region, and upper extremity in the primary motor cortex. For clarity, only one direction of the reciprocal sensorimotor pallidosubthalamic projection is shown. Note that the cohorts’ datapoints provide comprehensive coverage of the pathway activation space.

Fig. 4, A shows activation profiles of the best responders, i.e., patients with the highest UPRDS improvement from baseline, in both cohorts and the theoretical 100% improvement profile, derived from datapoints of three patients (from the training cohort) with the highest normalized UPDRS-III improvement. The Canberra distance of the datapoints from the rest of the training cohort to the optimal profile showed a significant inverse correlation with the correponding UPDRS-III improvements from baseline (see Fig. 5, A). In contrast, the optimal profile derived from all datapoints of the training cohort was not predictive (see Suppl. 2).

**Figure 4.**
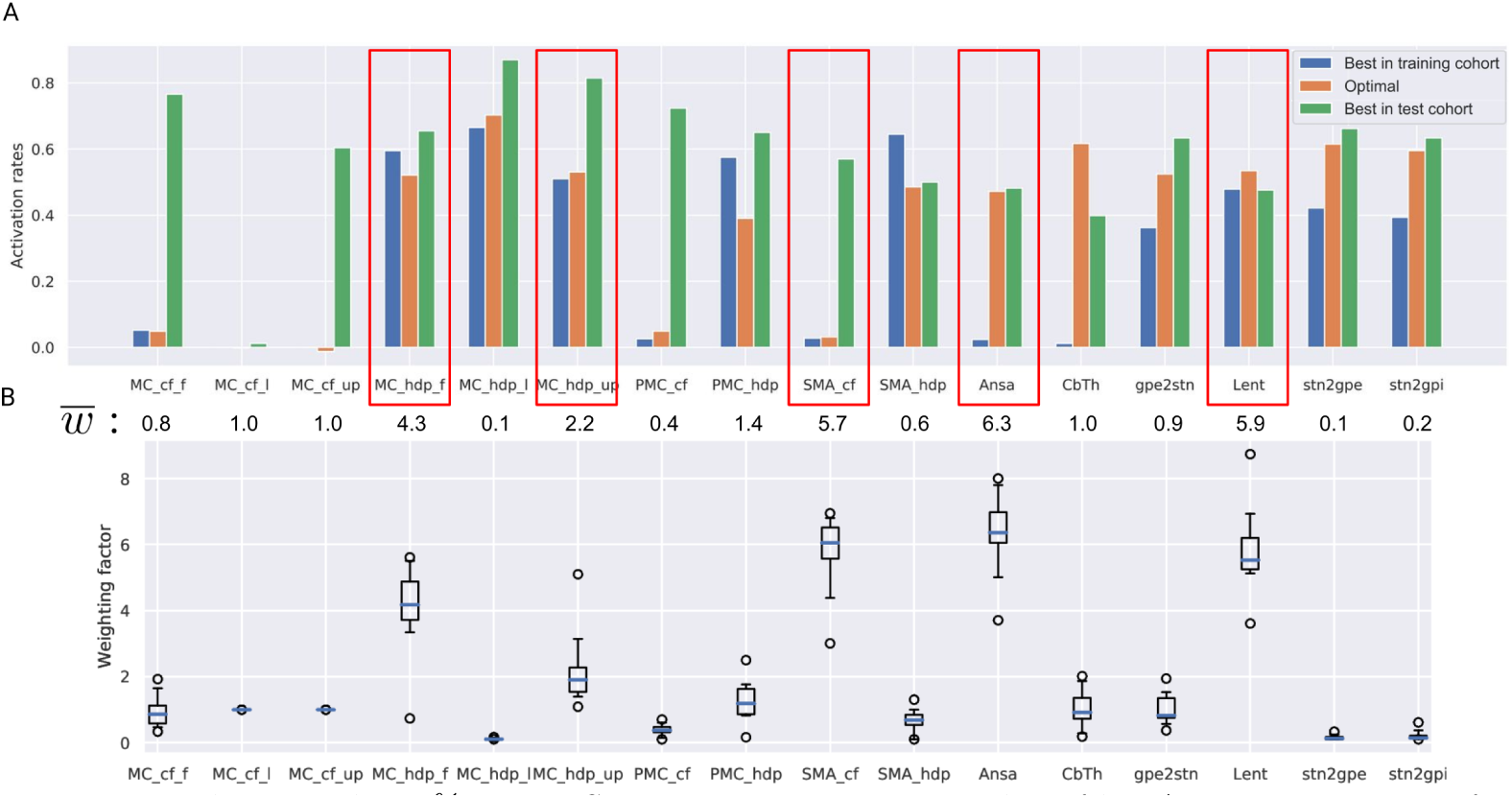
Theoretical 100% UPDRS-III improvement optimal profile. A: activation rates for the optimal profile and the best responders from each cohort. Note that the match between these datapoints and the optimal profile occurs for different pathways, suggesting that two distinct DBS mechanisms might be present. The mean values of optimized weighting factors below the bars can be interpreted as the importance of matching the optimal profile for a particular pathway. Red boxes highlight pathways with the highest effect on predictive ability. B: box plots of the optimized weighting factors computed with the ’leave-one-out’ approach. Note the low number of outliers.

**Figure 5.**
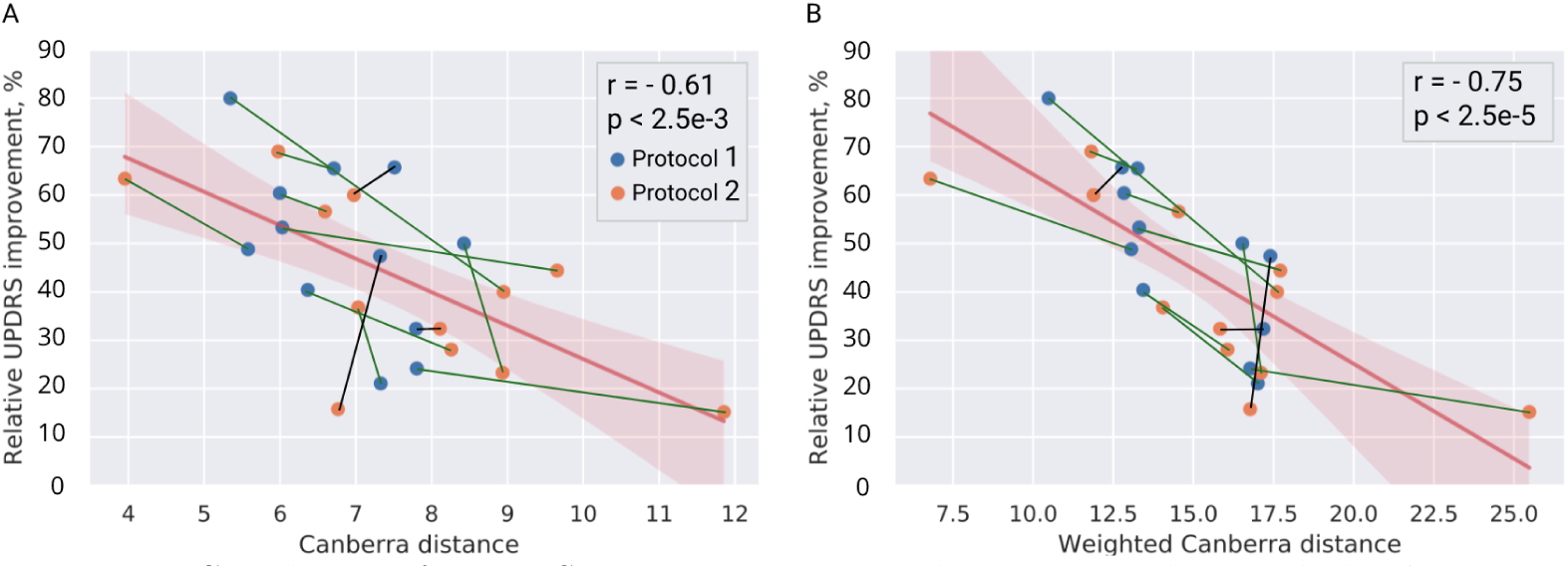
Correlation of UPDRS-III improvement in the training cohort with the distance to the optimal profile defined in the pathway activation space and based on three patients with the highest normalized interprotocol UPDRS-III improvement. Colored lines between datapoints depict interprotocol binary correlation (green – valid prediction, black – false.)

### Key Pathways of the Activation Profile

Optimization results for the weighting factors *w*_*i*_ in the Canberra distance (see Eq. 3), conducted on datapoints of the training cohort, indicated that DBS-induced activation in specific pathways might play a prominent role in motor symptom alleviation (see Fig. 4, B).

In particular, the optimization emphasized a moderate activation in both pallidothalamic pathways as well as two branches of the HDP to the primary motor cortex, while avoiding stimulation of the corticofugal pathway descending from the supplementary motor area. The weighted Canberra distances from activation profiles to **AR**_optimal_ for patients who were successively left out showed a statistically significant correlation with UPDRS-III improvement from baseline. Notably, the optimized weights were comparable among patients and were averaged to draw a general conclusion on the importance of particular pathways, where deviations in activation rates from the optimal profile led to worse performance.

The averagely weighted Canberra distance to **AR**_optimal_ demonstrated a higher correlation with UPDRS-III improvement from baseline than the unweighted metric (Fig. 5, B), which is not, however, surprising, since the optimization was conducted on these datapoints. Therefore, the profile-based predictive model had to be validated on unseen patients from the hold-out test cohort. Critically, patients from the test cohort did not play a role in the model selection process and were entirely naive to the final metric derived from the training cohort. This metric, the weighted Canberra distance to **AR**_optimal_, defined using the training cohort, was predictive of UPDRS-III improvement from baseline in patients of the test cohort, while the unweighted distance was not (see Fig. 6.) Distribution of activation in the highly weighted pathways computed for the best responders is shown in Fig. 7.

**Figure 6.**
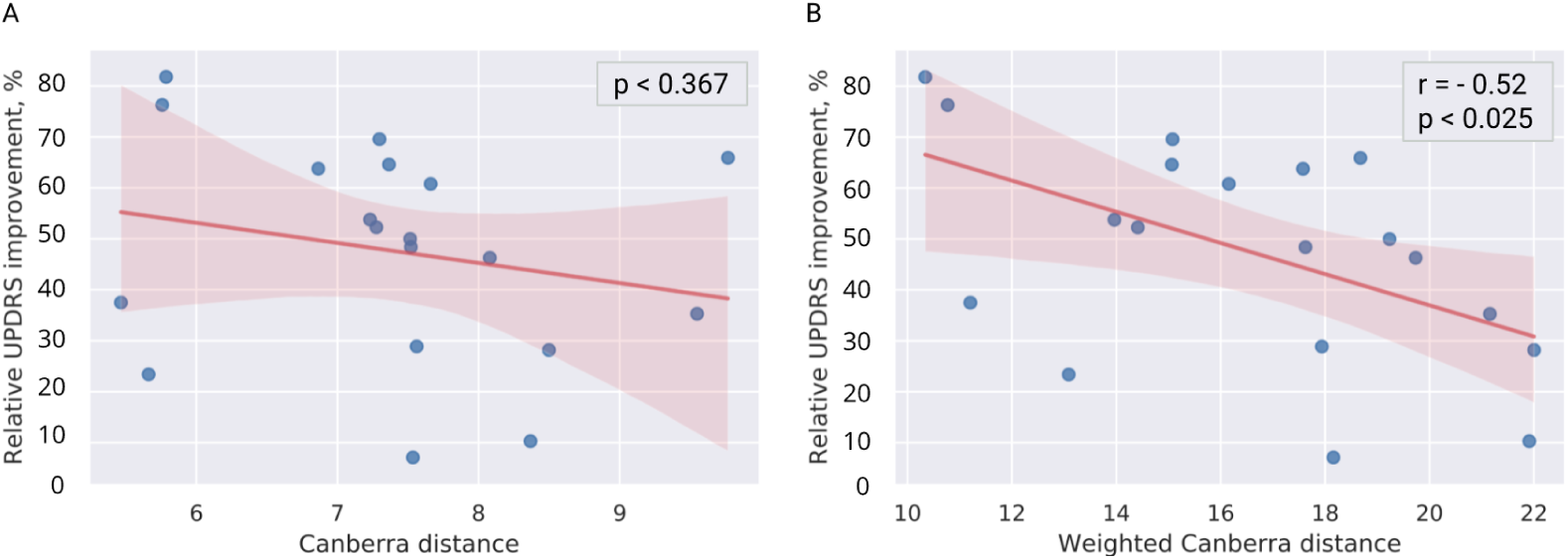
Cross-predicting UPDRS-III improvement in the test cohort using the optimal activation rate profile derived from datapoints of the three patients in the training cohort. Note that only the weighted Canberra distance has a statistically significant correlation with the improvement.

**Figure 7.**
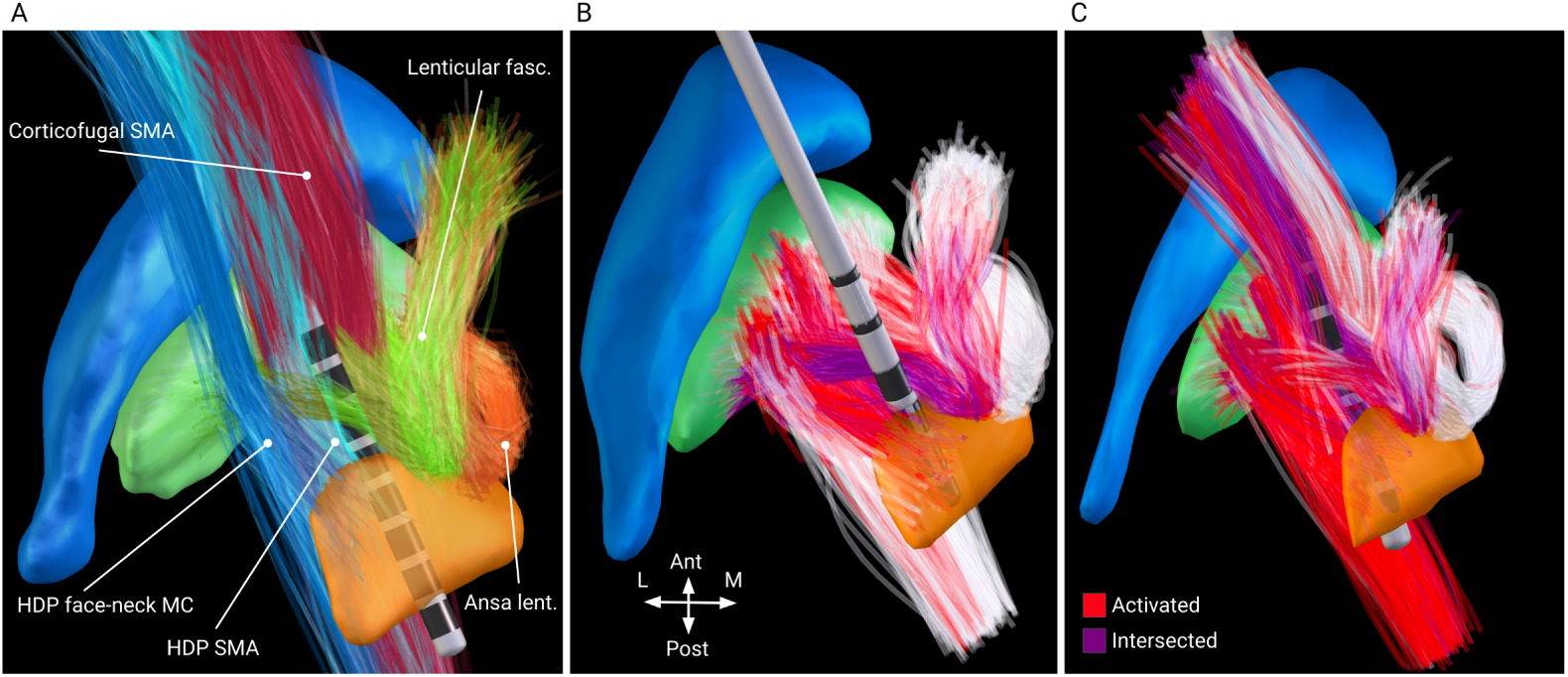
Pathway activation visualized in Lead-DBS. A: pathways which activation rates determine the UPDRS-III improvement (shown in MNI space.) Blue and cyan – the hyperdirect pathway, descending from face-neck and upper extremity of the primary motor cortex, red – the corticofugal pathway, descending from the supplementary motor area, green – lenticular fasciculus, orange – ansa lenticularis. B and C: pathway activation (shown on computational axon models in patient-specific space) for the best responders in the training and the test cohorts, respectively. Axons closer than 0.1 mm from the electrode (purple) are excluded due to glial scaring and neurodegeneration. Note the high recruitment of the HDP and the corticofugal pathway for the best responder in the test cohort.

## Discussion

In this study, we conducted pathway activation modeling for two cohorts of Parkinson’s patients that underwent DBS surgery at two independent centers. Based on modeling results for a training cohort, we proposed a method to derive an optimal activation profile using differences in activation rates and UPDRS-III scores assessed for two protocols evaluated in each patient. Furthermore, we evaluated the importance of activation in particular pathways for the obtained profile. At last, we demonstrated that proximity to the optimal activation profile in the multidimensional pathway activation space has a statistically significant correlation with UPDRS-III improvement in a test cohort from an independent center.

### Effect of Activation in Individual Pathways

Contrary to our prior expectations, the computed activation rates of individual axonal pathways demonstrated no clear correlation with clinical improvements. This result is especially remarkable for the hyperdirect pathway (HDP), associated with motor symptom alleviation in Parkinson’s disease (PD) [9, 39, 48]. Furthermore, even though activation in one branch of the HDP correlated with UPDRS-III improvement from baseline, the result was not reproducible in the test cohort. One possible explanation is that the correlation of improvement with HDP activation might be hindered by the nearby corticofugal pathway, whose stimulaton has been associated with tetanic motor contractions [51]. In addition, subthreshold stimulation of these tracts has been reported to exacerbate akinesia and bradykinesia [55], so patients may not necessarily show significant relief of motor symptoms despite high activation rates in the HDP. Admittedly, this was not the case for the best responder in the test cohort (see Fig. 3.), possibly due to its individual lower responsiveness to the stimulation. We also analyzed correlations of UPDRS-III improvement with activation in the HDP, the corticofugal, and the pallidothalamic pathways without differentiating between branches. However, this did not yield conclusive results. Although high activation in the corticofugal pathway induces motor contractions, such stimulation protocols are avoided in chronic DBS, so this side effect could not be quantified in the correlation tests.

For the cerebellothalamic pathway, we did observe a correlation between activation rates and tremor suppression, which is in agreement with other studies [2, 4, 10]. However, the correlation was present only when comparing two DBS protocols, but not DBS against baseline (see Suppl. 1). This suggests involvement of other pathways in thesuppression of tremors [52]. Furthermore, our model did not assign a high weighting of the cerebellothalamic pathway for predicting optimal outcomes. Nevertheless, the correlation of interprotocol differences implies that more effective alleviation of tremor could be achieved by steering current to the cerebellothalamic pathway.

### Effect of Balanced Pathway Activation

More conclusive results were obtained when considering motor improvement as a function of the whole activation profile. Defined by the weighted Canberra distance, discrepancy of test cohort profiles with the optimal activation rate profile demonstrated a statistically significant inverse correlation with UPRDS-III improvement from baseline. Importantly, the optimal profile was derived from datapoints of the training cohort, in particular, from differences of DBS-on intrapatient UPDRS-III scores and corresponding activation rates.

When analyzing the optimal profile, it is notable that two branches of the HDP to the primary motor cortex and of the corticofugal pathway to the supplementary motor area were assigned high weighting factors (see Fig. 4, B). For the latter, the activation rate in the optimal profile was minimal, suggesting its detrimental effect. On the other hand, the moderate activation rates in the presumably beneficial HDP branches can be explained by the proximity to the corticofugal pathway (note the high activation rates of the HDP and the corticofugal pathway for the best responder in the test cohort.)

The activation rates and the weighting factors for the optimal profile could be used to draw conclusions regarding possible mechanisms of action of STN-DBS. The most noteworthy is the presumed importance of the pallidothalamic projections: the ansa lenticularis and lenticular fasciculus, which have been considered a functional continuum [45, 46]. By comparing the weighted optimal profile to the best responders, we can hypothesize that these pathways as well as the HDP are beneficial for alleviation of motor symptoms in PD. The ansa lenticularis and lenticular fasciculus are the primary inhibitory outputs of the basal ganglia to the ventral anterior thalamic nucleus [37], which itself is projecting to motor-relevant cortical regions. In a PD affected network, the GPi exhibits bursting activity [50], which has been associated with generation of motor symptoms [35]. Alleviation could arise from a direct modulation of this pathological activity, e.g. via an “informational lesion” induced by the non-physiological pattern of DBS [21]. Furthermore, therapeutic effects of stimulation of the pallidothalamic pathways would also explain the comparable efficiency of GPi-DBS in treating motor symptoms of PD. In historical context of ablative surgery, lesioning of ansa lenticularis was shown to be beneficial for tremor alleviation [54].

Considering these pathways as potential targets has important clinical implications. For DBS patients who respond poorly to the treatment, such as those in whom the active electrode contacts are outside the dorsolateral “sweet spot” of the STN [11], clinicians could attempt to alleviate symptoms by thoroughly investigating stimulation responses for electrode contacts near the ansa lenticularis and lenticular fasciculus. Prospective studies could consider new implantation trajectories that allow both mechanisms with precise targeting of the hyperdirect and pallidothalamic pathways, preferably stimulated via different electrode contacts. This strategy would offer the possibility of inducing desynchronised DBS patterns.

At the same time, one should be aware that the STN is a site of convergence of different functional circuits [1], which is the most probable reason for its efficiency as a DBS target. Apart from the obvious cases, such as the near-zero activation of the corticofugal branch to the supplementary motor area (Fig. 4), the weighted profile of optimal activation rates does not suggest which pathways are beneficial or detrimental, but rather indicates a theoretically optimal balance of DBS-induced activation. Such a metric is more feasible than separately defined optimal activation rates, especially since pathway-specific DBS is not yet possible with clinically approved electrodes.

### Limitations

The most disputable result presented in this study is the high activation in the corticofugal pathway computed for the best responder in the test cohort. For clinical application, this activation profile would be undesirable due to possible side effects. At the same time, the activation of the corticofugal branch to the lower extremity of the primary motor cortex was nearly zero for this patient, as well as for others in both cohorts (see Fig. 3). Further studies are warranted to investigate pathway activation profiles associated with DBS-induced side-effects.

Nevertheless, one should consider that the arguable results of correlation tests for individual pathways might originate from uncertainties introduced by the computational model. We suspect that the lack of data on pathway-specific axonal morphology is the primary source of the error in pathway activation modeling. Furthermore, additional inaccuracies arise from volume conductor modeling, processing of medical images and clinical evaluation. Investigating present results by means of uncertainty quantification, as was shown in [6, 49] in the context of DBS, could enhance our understanding of how results should be interpreted.

In the optimal profile, noteworthy is the small weighting factor of the sensorimotor pallidosubthalamic and reverse projections. That might be attributed to the limitations of the pathway activation modeling when investigating this local circuit, which excitatory-inhibitory reciprocity is a probable source of synchronization in the basal ganglia [44]. These short projections follow nearly the same trajectory, hence nearly the same DBS-induced activation rates, and a neural activity of one pathway is directly modulated not only by the DBS-induced membrane polarization, but simultaneously via the activity of the reciprocal pathway, which is also recruited by DBS. Therefore, a simulation model that could more accurately represent the circuit would need to account for temporal dynamics and consider effects of extracellular stimulation on synaptic inputs [12]. Besides, due to the electrode implantation, these short pathways are relatively more exposed to glial scaring that potentially affects neurotransmission and extracellular ionic concentrations [33].

Apart from the limitations of the computational model mentioned above, the following study limitations deserve mentioning. First, not all pathways affected by STN-DBS were modeled. To decrease the computational effort, we deliberately excluded the anterior cingulate and the prefrontal cortex assuming that they play a minor role in the motor symptoms of Parkinson’s disease, although connectivity with the latter was associated with rigidity improvement [3]. Among the local projections not examined, pathways of the substantia nigra pars reticulata are of particular interest, as it plays a similar role in the basal ganglia as the GPi. Unfortunately, these projections were not included in the basal ganglia pathway atlas [47], employed here as an exclusive source of the structural connectivity.

Developed by experienced neuroanatomists using advanced visualization techniques, this atlas is less prone to contain false-positive trajectories occurring due to a poor signal-to-noise ratio of diffusion imaging, commonly employed for individualized fiber tractography. However, the atlas is neither patient-specific nor able to account for disease-related changes. In the present study, individual anatomical variability was partially accounted for by translation of pathways to the patient-specific space using a subcortical normalization strategy [14].

It should be noted that the optimal profile was derived based on UPDRS-III improvement in the training cohort, while validated on the test cohort assessed with MDS-UPDRS-III score. However, the scores are strongly correlated (*r >* 0.95) [42] and were shown to be linearly dependent [19, 25], hence invariant in the Pearson correlation test. Furthermore, to generalize UPDRS-III improvement, we avoided differentiating between lateral and axial symptoms when analyzing activation rates, which were calculated separately for each hemisphere and then averaged. Differences in the pathway activation rates across both hemispheres are presented in Suppl. 3.

Finally, the optimization procedure of the weighting factors, conducted in the training cohort, was developed rather intuitively, and future studies should consider more robust methods. It also remains unclear whether optimization should emphasize the interprotocol correlation of the improvement with the distance to the optimal profile or a general increase of correlation for all datapoints. For the given datapoints, we did not observe significantly higher predictability in the test cohort for either case.

## Data Availability

The data that support the findings of this study are available from the first corresponding author.

