## Supplementary material for "Linking Profiles of Pathway Activation with Clinical Motor Improvements – a Retrospective Computational Study": Suppl. material

***Suppl. 1***

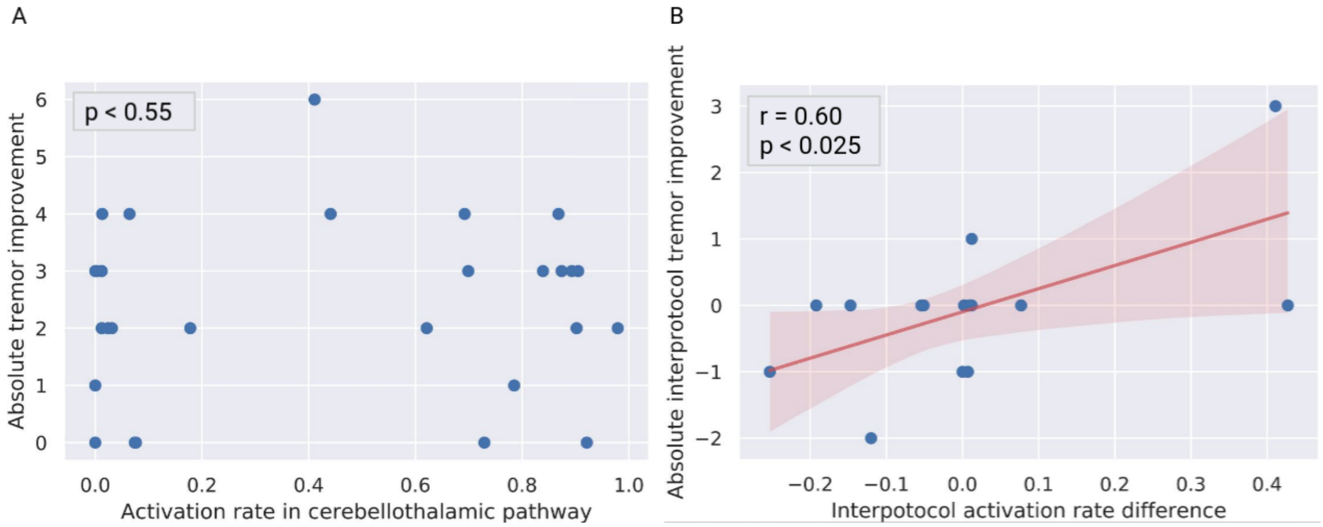


*Effect of activation in the cerebellothalamic pathway on tremor alleviation. A: there was no correlation observed between activation rates and tremor improvement from baseline. B: interprotocol difference in activation rates correlated with the corresponding difference in the tremor scores.*

***Suppl. 2***

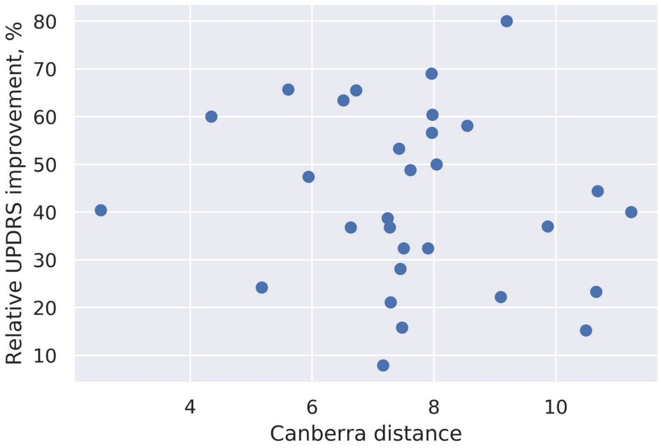


*Absence of correlation of UPDRS improvement with the distance to the target profile defined in the pathway activation space based on all patients of the training cohort.*

***Suppl. 3***

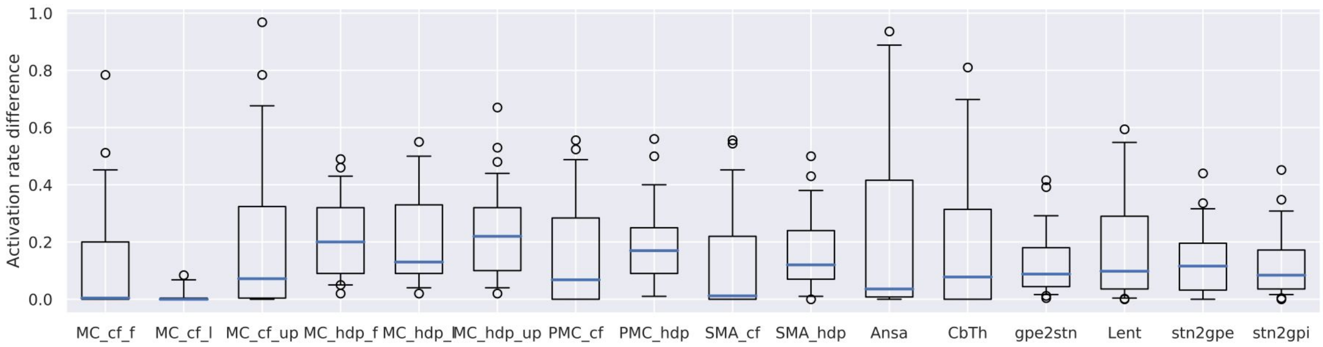


*Box plot of absolute difference of activation rates computed separately for both hemispheres across both cohorts. MC and PMC refer to the primary and premotor cortical regions, SMA -- supplementary motor area; cf and hdp are the corticofugal and the hyperdirect pathways, respectively; Ansa -- ansa lenticularis, Lent -- lenticular fasciculus, CbTh -- cerebellothalamic pathway; l, f, up - lower extremity, face-neck region, and upper extremity in the primary motor cortex.*
